# Carriage duration of carbapenemase-producing *Enterobacteriaceae* in a hospital cohort - implications for infection control measures

**DOI:** 10.1101/19001479

**Authors:** Yin Mo, Anastasia Hernandez-Koutoucheva, Patrick Musicha, Denis Bertrand, David Lye, Ng Oon Tek, Shannon N. Fenlon, Swaine L. Chen, Ling Moi Lin, Wen Ying Tang, Timothy Barkham, Niranjan Nagarajan, Ben S Cooper, Kalisvar Marimuthu

## Abstract

Carriage duration of carbapenemase-producing *Enterobacteriaceae* (CPE) is uncertain. We followed 21 CPE carriers over one year. Mean carriage duration was 86 (95%CrI= [60, 122]) days, with 98.5% (95%CrI= [95.0, 99.8]) probability of decolonization in one year. Antibiotic consumption was associated with prolonged carriage. CPE-carriers’ status should be reviewed yearly.

## Introduction

The rapid global dissemination of carbapenemase-producing *Enterobacteriaceae* (CPE) poses a public health threat [1]. The lack of effective therapy is associated with high mortality and healthcare costs [1]. Infection prevention and control are important measures to prevent the spread of CPE to vulnerable patients.

Early identification, isolation, and contact precautions are advocated by international guidelines to prevent the spread of CPE in healthcare settings [2,3]. Understanding the natural history and duration of CPE carriage will help in the design of rational infection control policies. In this study, we estimate carriage duration of CPE in a hospital cohort and identify risk factors for prolonged carriage.

## Methods

We conducted a prospective cohort study involving CPE carriers from October 2016 to February 2018. Study participants were recruited from two tertiary centers with over 1600 beds in Singapore. All admitted patients above 21 years with prior hospitalization within 12 months were screened for CPE carriage. We collected stool samples from study participants on enrolment, weekly for four weeks, monthly for five months, then bimonthly for six months. Demographic characteristics, healthcare contact and medication history were recorded. The study received ethics approval from the Singapore National Healthcare Group Domain Specific Review Board (NHG DSRB Reference: 2016/00364) prior to commencement.

Stool samples were inoculated onto selective chromogenic agar (ChromID Carba SMART), and species identification was done with matrix assisted laser desorption ionization-time of flight-mass spectrometry (MALDI-ToF (Bruker)). Antibiotic susceptibility testing was performed using VITEK-2. All *Enterobacteriaceae* isolates with a minimum inhibitory concentration of ≥2mg/L for meropenem, or ≥1.0mg/L for ertapenem, underwent polymerase chain reaction (PCR) for the presence of *bla*_NDM-1_, *bla*_KPC_, *bla*_OXA-48_, *bla*_IMI-1_ and *bla*_IMP_ genes [4]. All CPE isolates and stool DNA underwent sequencing on the Illumina HiSeq 4000 sequencer. The Shannon diversity index was used to measure α-diversity for stool microbial communities [5].

We analyzed the data with Bayesian multi-state Markov models to account for interval censoring (Supplementary material). First, we estimated the overall transmission rates by considering patients to be in either CPE colonized or non-colonized states. Secondly, we considered CPE colonization on a species level, and included CP-*Escherichia coli* (CP-EC) colonized, CP-*Klebsiella pneumoniae* (CP-KP) colonized, CP-EC/KP co-colonized as separate states (Supplementary figure 1). All analyses were performed using R 3.4.4 and RStan [6,7].

## Results

Twenty-one patients were enrolled with a mean follow-up period of 294 (SD=77) days, and each participant provided 12 (SD=1.5) samples. Patient details on demographics and medical history are in Table 1.

**Table 1.**
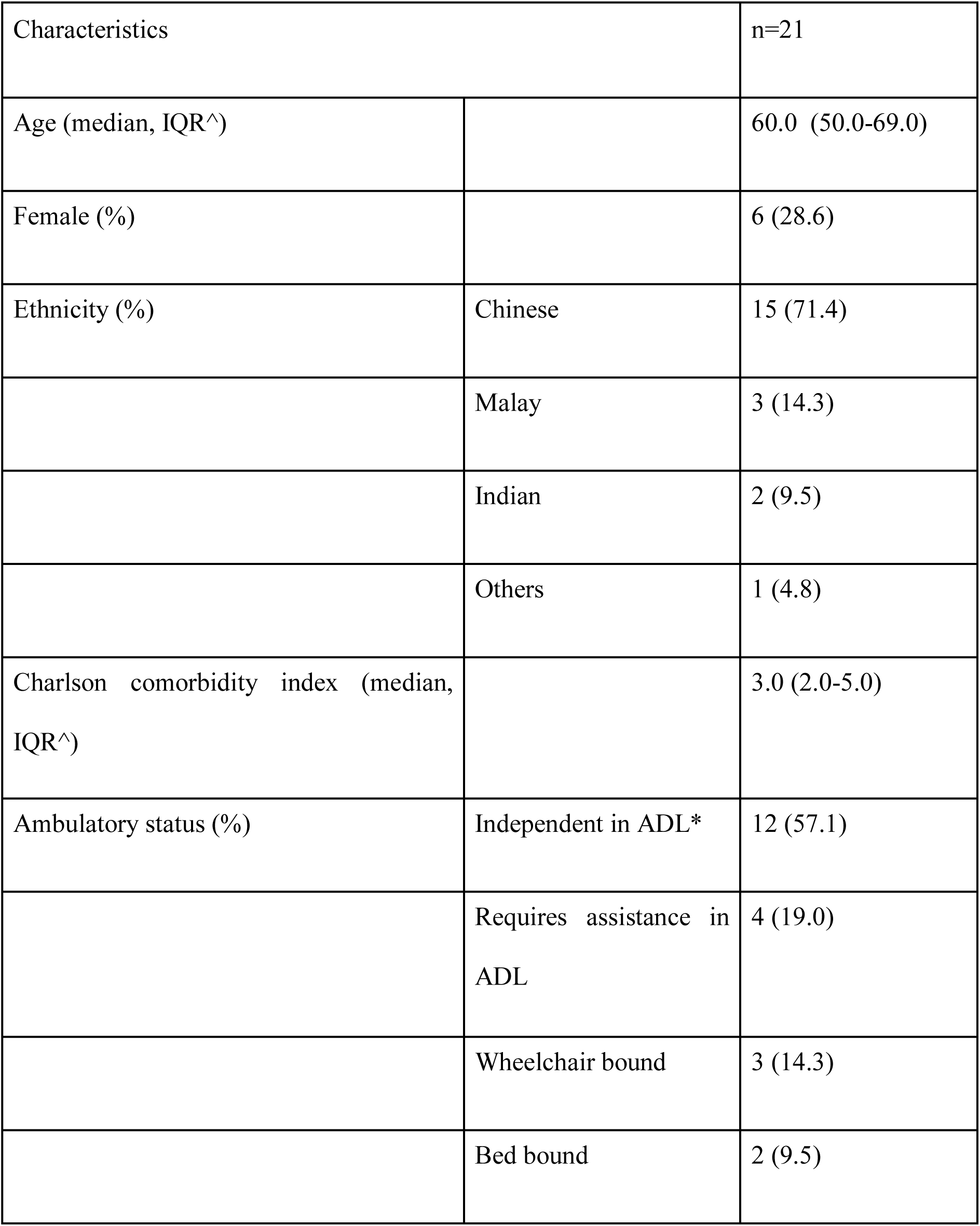
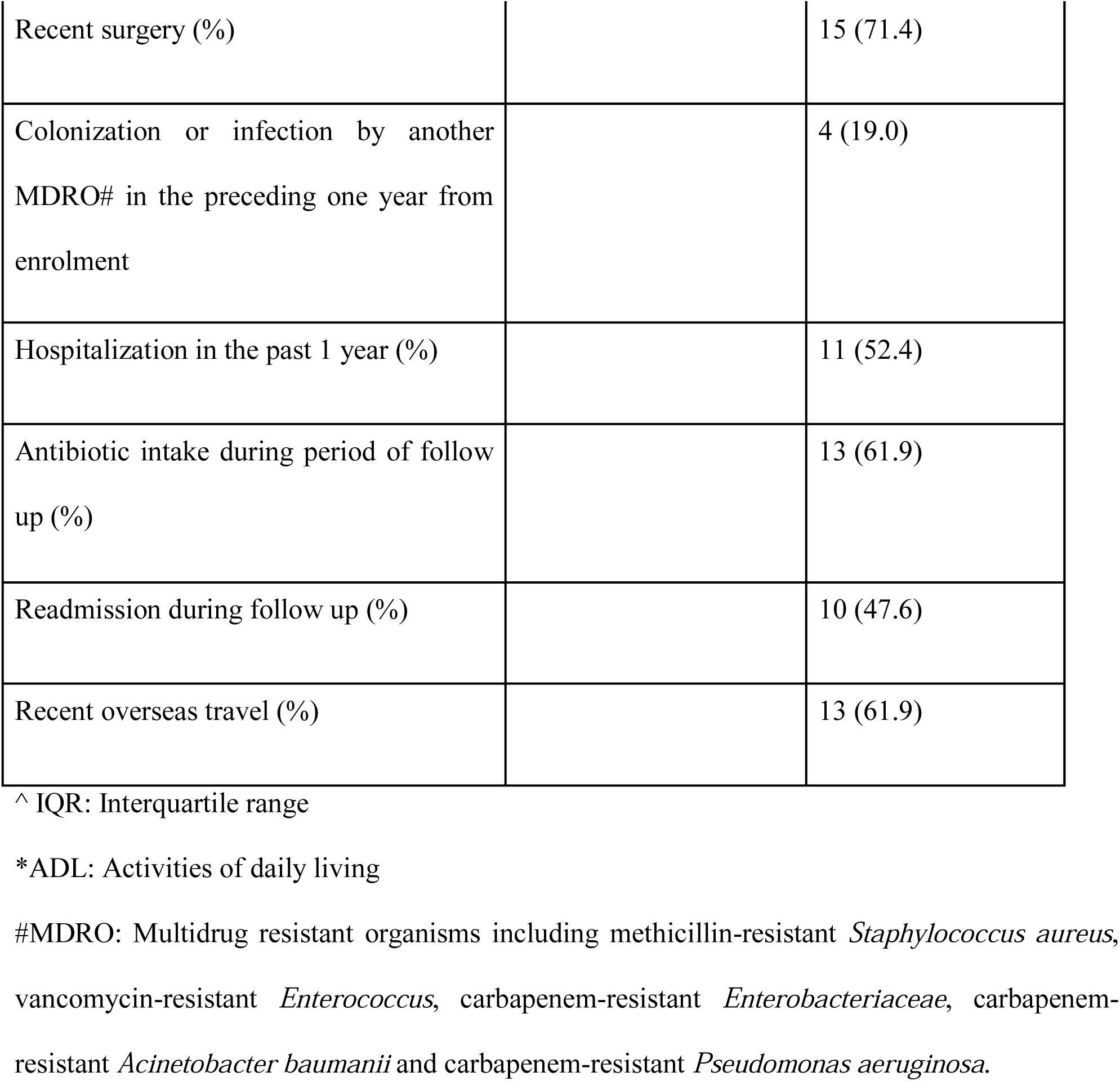
Participant demographics

Throughout follow-up, 15/21 (71.4%) participants carried more than one species of CPE, while only 3/21 (14.3%) participants carried more than one type of carbapenemase genes (χ^2^ of difference in proportions=14.8, simulated p-value=0.0005) (Supplementary table 1). The most common species carried by the participants were *K. pneumonia*e (18/21, 85.7%) and *E. coli* (16/21, 76.2%). The most frequently observed carbapenemase genes were OXA-48 (11/21, 52.4%) and KPC (8/21, 38.1%). Seventy-six CP-KP were isolated from the samples, out of which sequence type (ST) 307 (25/76, 32.9%) was the most common. Amongst the CP-EC isolates, ST131 (22/83, 26.5%) was the most common. Most participants (17/21, 81.0%) had continuously positive samples until clearance. Four participants had negative samples followed by positive samples, with the longest period being three negative samples over three consecutive weeks (Figure 1).

**Figure 1.**
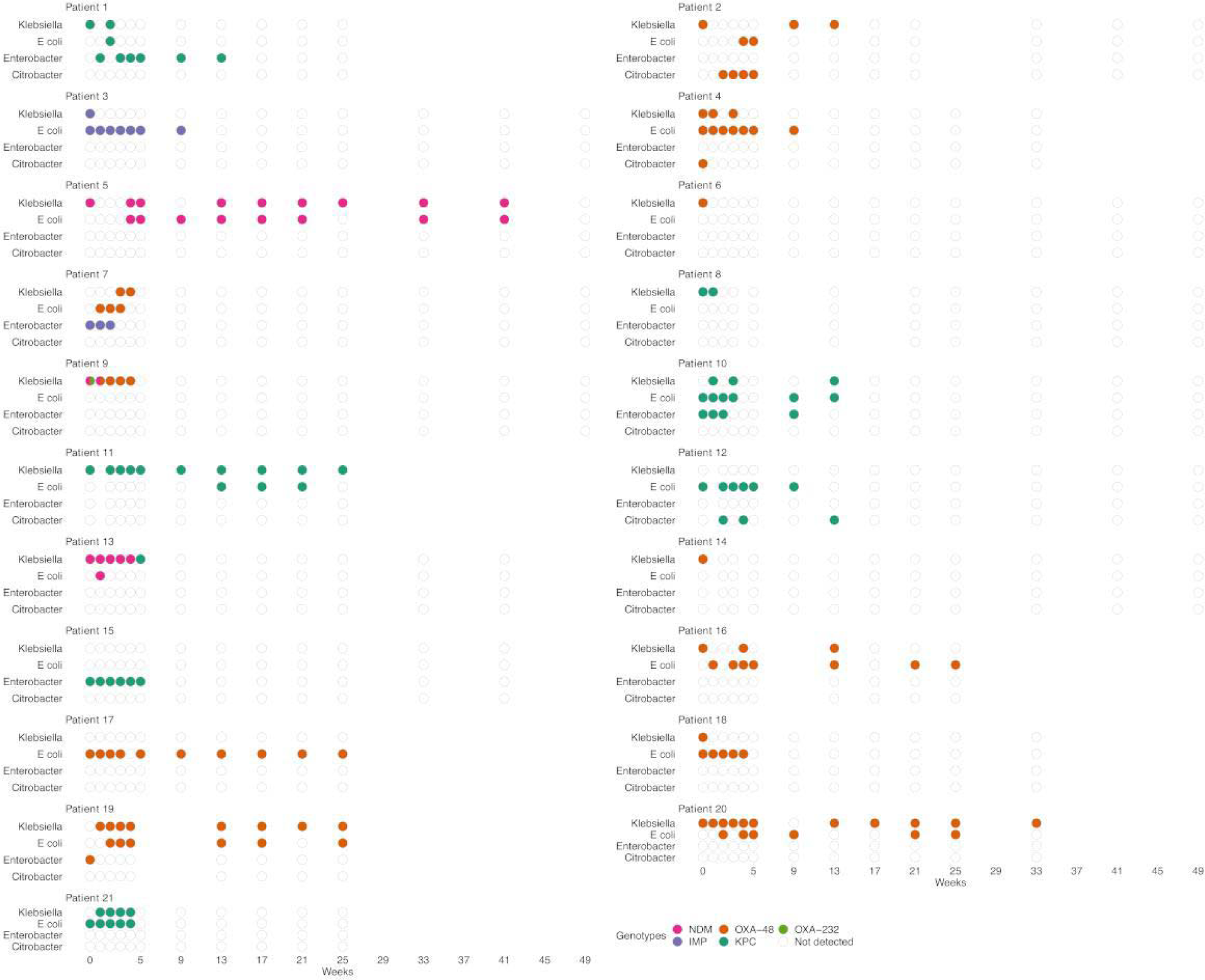
Types of CPE and plasmid colonization for study participants. Figure 1 depicts the microbiological outcomes of stool samples for each participant. Each column of dots represents one stool sample. Each dot represents an *Enterobacteriaceae* species, including *Klebsiella spp*., *E coli, Enterobacter spp. and Citrobacter spp*., yielded in the stool samples carrying CPE genes. Missing data is shown by the absence of dots on a particular week.

The estimated mean duration of CPE carriage was 86 (95%CrI= [60, 122]) days. The probability of decolonization was 98.5% (95%CrI= [95.0, 99.8]) in one year, assuming a constant decolonization rate within the time interval. The longest observed carriage duration was 387 days. We analyzed age, co-colonization with other multidrug resistant organisms, presence of urinary catheter, antibiotic use during follow up, Charlson comorbidity index, re-admission, and Shannon diversity index as covariates to explore their association with decolonization (Supplementary Figure 2). Antibiotic use during follow up was the only factor associated with prolonged CPE carriage (HR= 0.48, 95%CrI= [0.20,0.93]). The rate of decolonization from CP-EC was lower than that of CP-KP (0.018/day, 95%CI=[0.007,0.031] versus 0.030/day, 95%CI=[0.016, 0.049]) (Supplementary table 2 and figure 3).

## Discussion

CPE infections are typically preceded by asymptomatic carriage, especially in vulnerable patients such as the immunocompromised and critically ill [8]. Active surveillance to identify CPE carriers is essential to prevent transmissions, but may be associated with high cost-benefit ratio if implemented without understanding the natural history of CPE carriage.

Previous reports of CPE carriage duration are highly varied, with median duration ranging from 43 days to 295 days[8,9]. This is likely due to different follow-up schedules, microbiological and molecular methods used to identify CPE, and criteria to define clearance. Studies that reported longer carriage duration tended to adopt an opportunistic sampling strategy, and considered both clinical and stool samples to determine carriage [9]. Opportunistic sampling may lead to selection bias as patients with more healthcare contacts would have more samples taken. Infrequent and inconsistent sampling is more likely to misclassify re-colonization from a new transmission event as continuous colonization, resulting in perceived longer duration of carriage.

Our systematic sampling and robust methods to identify CPE allowed us to closely follow the participants’ carriage status. With combined detection methods of culture on carbapenem-resistance selective media, antibiotic susceptibility testing and PCR, we found that a proportion of patients (4/21, 19.0%) had intervening negative samples, ranging from one to three weeks apart. To confidently declare eradication, a patient should have at least two negative samples four to six weeks apart. Given that the probability of decolonization is 98.5% in one year, we suggest that CPE carriers status should be maintained for one year. Further health economics analysis is needed to make institution-specific recommendations on frequency of re-screening. Given the finding that antibiotic use was the most important factor associated with prolonged CPE carriage, antibiotics should be avoided if not clinically indicated.

An interesting finding is that the participants carried more species of CPE than types of carbapenemase genes. While this may be due to new acquisition events, this observation is more parsimoniously explained by active interspecies horizontal gene transfer especially in a low-transmission setting such as in Singapore. Differential rate of clearance of CP-KP and CP-EC can be related to colonizing affinity of the species and fitness cost of carbapenemase genes, which are highly variable amongst different species [10]. Further studies incorporating both between and within host transmission dynamics of resistance will be important to shed light on the roles of bacterial clones and plasmids in spreading and maintaining resistance.

A limitation of the study is that the participants were screened after hospital admission and the time of initial colonization could not be confidently determined. However, a recent survey of the Singapore community showed that none out of 305 community dwellers carried CPE in their stools [11]. Therefore, the likelihood that the participants were already colonized by CPE in the community is low. Though our sample size is relatively small, to our knowledge, this study is the most rigorously conducted in terms of frequency of stool sample collection, duration of follow up, and number of participants in a non-outbreak setting. The use of multistate models has been shown to preserve power with modest sample size given more frequent follow-ups [12].

## Role of funding

The study is primarily supported by the Singapore Biomedical Research Council-Economic Development Board Industry alignment fund (Grant ref: IAF311018). MY is supported by the Singapore National Medical Research Council Research Fellowship (Grant ref: NMRC/Fellowship/0051/2017). BSC is supported by UK Medical Research Council / Department for International Development (Grant ref: MR/K006924/1). PM is supported by the JPI-AMR (MODERN) (Grant ref: C-17-0014). AH is supported by Wellcome Trust (Grant ref: 212630/Z/18/Z). The Mahidol-Oxford Tropical Medicine Research Unit is part of the Wellcome-Trust Major Overseas Programme in SE Asia (Grant ref: 106698/Z/14/Z). Additional grant support was provided by the NMRC Clinician-Scientist Individual Research Grant (Grant ref: NMRC/CIRG/1463/2016 and NMRC/CIRG/1467/2017), Singapore Ministry of Education Academic Research Fund Tier 2 grant: New Delhi Metallo-Beta-Lactamase: A global multi-centre, whole-genome study (Grant ref: MOE2015-T2-2-096), NMRC Collaborative Grant: Collaborative Solutions Targeting Antimicrobial Resistance Threats in Health Systems (CoSTAR-HS) (Grant ref: NMRC CGAug16C005), NMRC Clinician Scientist Award (Grant ref: NMRC/CSA-INV/0002/2016).

## Data Availability

Data is available on request to the corresponding author.

## Acknowledgement

All of the authors have no conflict of interest to declare.

## Supplementary material

Supplementary material 1: Methodology

### Microbiology

Microbiological cultures to identify CPE were performed via direct inoculation of stool samples onto selective and indicative agar, chromID® CARBA SMART Agar (CARB/OXA, Biomerieux). After overnight incubation, colonies were identified at the species level with matrix assisted laser desorption ionization-time of flight-mass spectrometry (MALDI-ToF-MS, Bruker Daltonics GmHB, Bremen, Germany). Phenotypic antimicrobial susceptibility testing was performed using VITEK-2 (bioMérieux Vitek, Inc., Hazelwood, MO). All *Enterobacteriaceae* isolates with a minimum inhibitory concentration to meropenem ≥2mg/L, or Ertapenem MIC ≥1.0mg/L, underwent polymerase chain reaction (PCR) to test for the presence of blaNDM-1, blaKPC (blaKPC-2 to blaKPC-13), blaOXA48, blaIMI-1 and blaIMP carbapenemase genes, as previously reported. (Teo *et al*, Singapore J Med Microbiol. 2013)

### Genomic analysis

Library preparation for DNA from CPE isolates was performed using the NEBNext® Ultra™ DNA Library Prep Kit for Illumina®. Sequencing with 2×151bp reads was performed using the Illumina HiSeq 4000 sequencer.

Raw FASTQ reads were processed using standard in-house pipelines. Briefly, MLST and antibiotic resistance genes were called directly from raw reads as well as from *de novo* assemblies; discrepancies between these were resolved with manual examination of both types of data. For analysis of raw reads, MLST and antibiotic resistance genes were called directly from the FASTQ files using SRST2 (version 0.1.8) (Inouye *et al*, Genome Med. 2014) with default settings using the ARGAnnot database provided with the SRST2 distribution for resistance genes. *De novo* assemblies were performed using the Velvet assembler (version 1.2.10) (Zerbino *et al*, Genome Res. 2008) with parameters optimized by the Velvet Optimiser script packaged with the velvet distribution, scaffolded with Opera (version 1.4.1) (Gao *et al*, J Comput Biol. 2011), and finished with FinIS (version 0.3) (Gao *et al*, Algorithms in Bioinformatics 2012). Genomes were annotated with Prokka (version 1.11) (Seemann *et al*, Bioinformatics. 2014). For analysis of *de novo* assemblies, resistance genes were called using BLASTN with a minimum identity of 90% over 90% of the gene length required for calling a gene present, using the same ARGAnnot database as used by SRST2. MLST calls were made by using a custom BLASTN-based MLST caller. The MLST databases were downloaded using the SRST2 helper scripts from https://pubmlst.org.

### Shotgun metagenomics

DNA from stool samples was extracted using PowerSoil® DNA Isolation Kit (12888, MOBIO Laboratories) with modifications to the manufacturer’s protocol. To avoid spin filter clogging, we extended the centrifugation to twice the original duration and solutions C2, C3 and C4 were doubled in volume. DNA was eluted in 80µL of Solution C6. Concentration of DNA was determined by Qubit dsDNA BR assay (Q32853, Thermo Fisher Scientific). For the library construction, 50ng of DNA was re-suspended in a total volume of 50µL and was sheared using Adaptive Focused Acoustics^™^ (Covaris) with the following parameters; Duty Factor: 30%, Peak Incident Power (PIP): 450, 200 cycles per burst, Treatment Time: 240s. Sheared DNA was cleaned up with 1.5× Agencourt AMPure XP beads (A63882, Beckman Coulter). Gene Read DNA Library I Core Kit (180434, Qiagen) was used for end-repair, A-addition and adapter ligation. Custom barcode adapters (HPLC purified, double stranded, 1^st^ strand: 5’ P-GATCGGAAGAGCACACGTCT; 2^nd^ strand: 5’ ACACTCTTTCCCTACACGACGCTCTTCCGATCT) were used in replacement of Gene Read Adapter I Set for library preparation. Library was cleaned up twice using 1.5× Agencourt AMPure XP beads (A63882, Beckman Coulter). Enrichment was carried out with indexed-primers according to an adapted protocol from Multiplexing Sample Preparation Oligonucleotide kit (Illumina). We polled the enriched libraries in equi-molarity and sequenced them on an Illumina HiSeq sequencing instrument to generate 2 × 101 bp reads, yielding 17,744 million paired-end reads in total and 49 million paired-end reads on average per library.

Reads were processed with an in-house shotgun metagenomics data analysis pipeline (https://github.com/CSB5/shotgun-metagenomics-pipeline). Read quality trimming was performed using famas (https://github.com/andreas-wilm/famas, v0.10, --no-order-check), and bacterial reads were identified by mapping to the human reference genome hg19 using bwa-mem (v0.7.9a, default parameters).

Microbial community taxonomic profiles were obtained using MetaPhlAn (v2.0, default parameters, relative abundance >0.01%) which provides relative abundances of bacteria, fungi and viruses at different taxonomic levels. The Shannon diversity index was computed from species-level taxonomic profiles using the function diversity from the R library vegan. The detection of antibiotic resistance genes was performed using SRST2 (v0.1.4, fraction of gene covered >99%) using the predefined ARGAnnot database.

**Supplementary Table 1:**
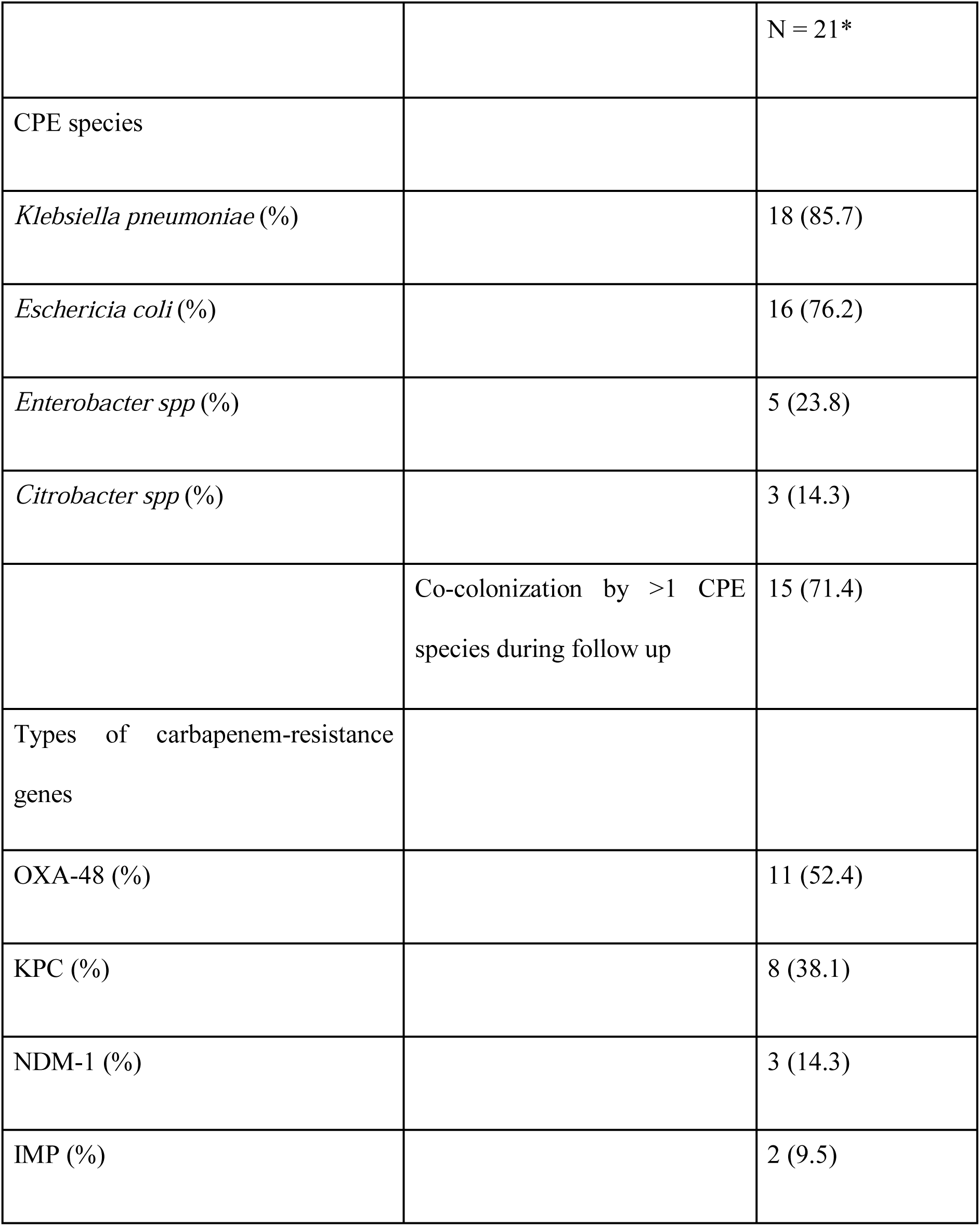
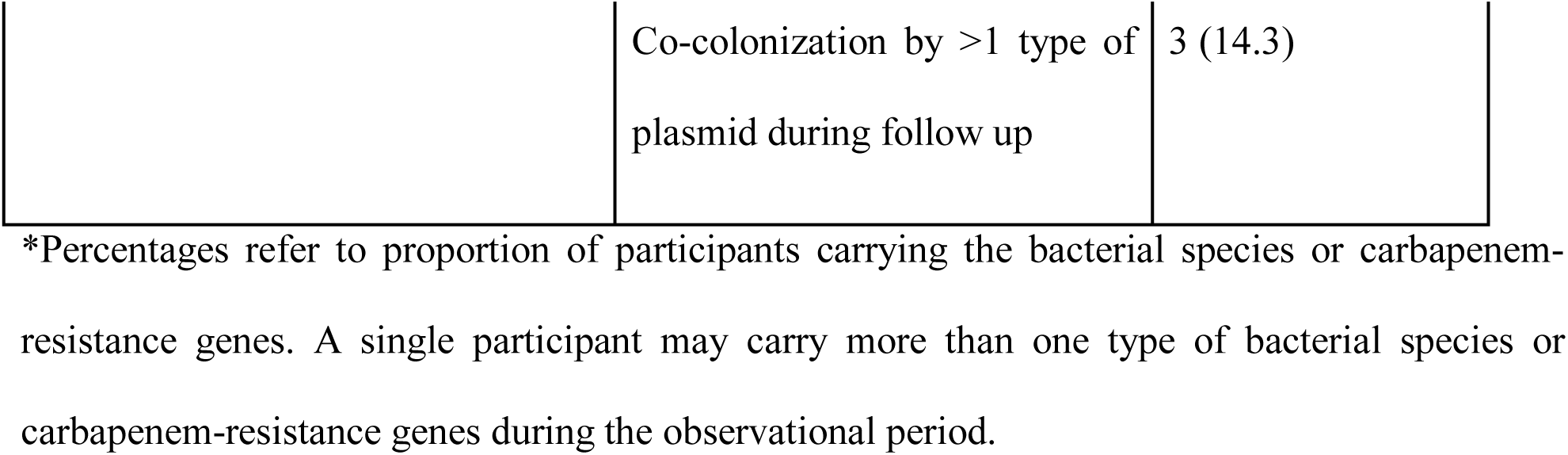
Carbapenamase producing *Enterobacteriaceae* and associated plasmids carried by the participants throughout the study period

**Supplementary Figure 1:**
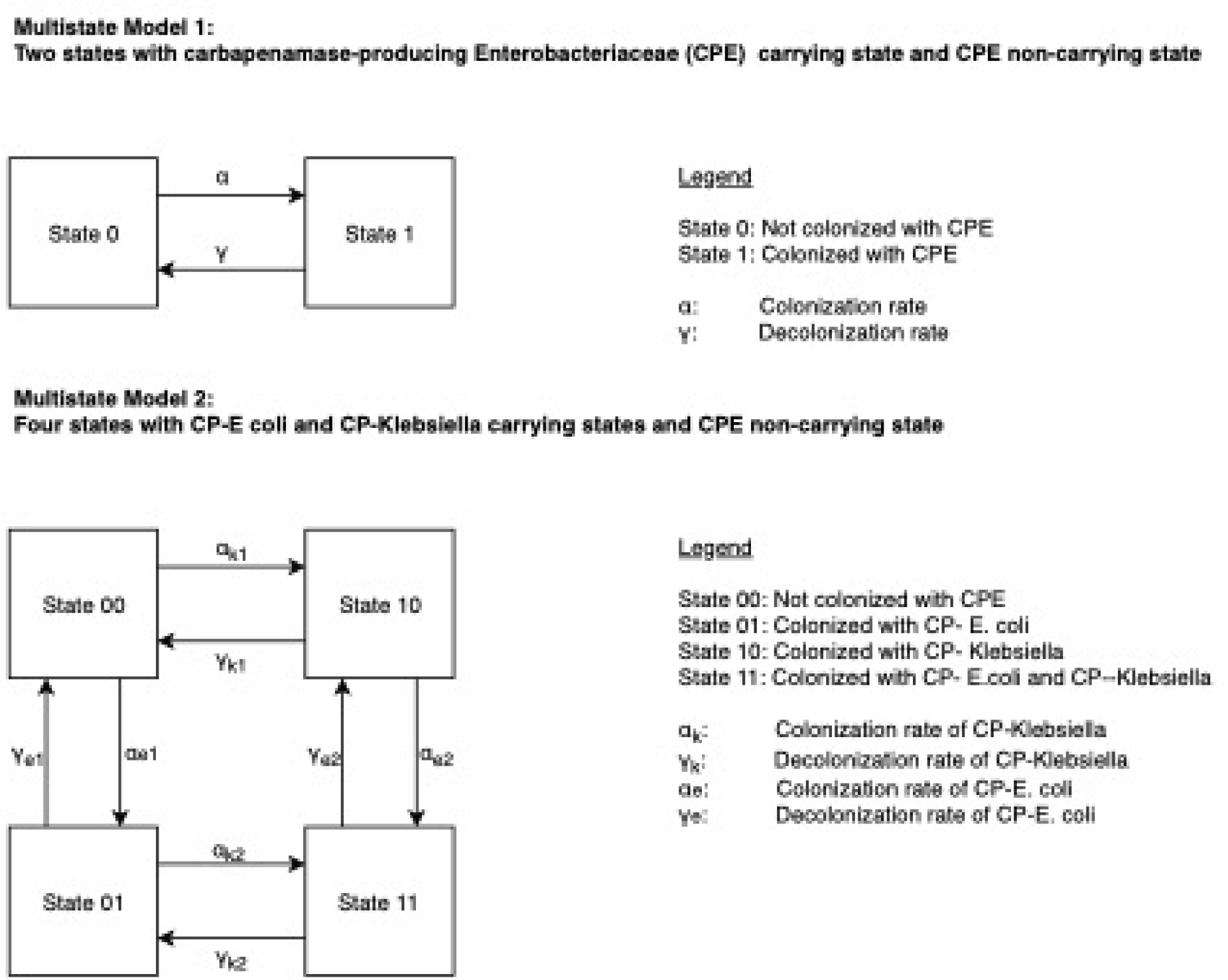
Multistate Markov models for the analysis of CPE carriage.

To estimate rates of colonization and duration of carriage, we modeled colonization and carriage dynamics using multi-state Markov models. First, we considered CPE to be a homogeneous bacterial group and at any sampling time point patients belonged to one of two states: “non-colonised” or “colonised “. Secondly, we considered CPE by species with patients being “not colonized by any CPE”, “colonized by CP-*E. coli*”, “colonized by CP-*K. pneumoniae*” or colonised by “both CP*-E. coli* and *K. pneumoniae*”, resulting in a four-state Markov model. In either models, transitions from one state to another was governed by a ***K*×*K*** intensity matrix Q. For ***r* ≠ *s***, the rate of transition from state r to state s, ***q***_***rs***_ **= *Q*[*r***,***s*]** was modelled by the linear equation:

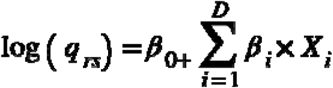

Where **{*X***_**1**_,***X***_**2**_, **≠ *X***_***D***_**}** is a set of covariates.

For the two-state homogeneous CPE modelled we included age, taking antibiotics during follow-up, Charlson comorbidity index, co-colonization with other multidrug resistant organisms, infection with CPE, readmission, Shannon diversity index and presence of urinary catheter as covariates.

The rate of remaining in state ***r*** was estimated by:

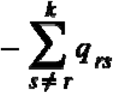

We implemented the models in STAN modelling language within the R environment. We used N∼(0,1) as prior distributions for parameters estimates, while posterior distributions were sampled using the Hamiltonian Markov Chain Monte Carlo method. The prior is a generic weakly informative prior as recommended by Rstan which contains enough information to regularize i.e. rule out unreasonable values but is not so narrow as to rule out probable values (Gelman, Github. 2019). Posterior distributions were sampled from 4 chains run over 20,000 iterations (including 10,000 burn-in) and we assessed model convergence using the Gelman-Rubin convergence diagnostic statistic and through visualization of trace plots.

**Supplementary Table 2:** Rates of the four-state multistate model with CP-*E. coli*, CP-*Klebsiella*, carrying both CP-*E. coli* and CP-*Klebsiella* (Both), CPE-non-carrying state (None)

|  | Event /day | 95% CrI |
| --- | --- | --- |
| CP-*E. coli* to None | 0.018 | 0.007 – 0.031 |
| CP-*Klebsiella* to None | 0.030 | 0.016 – 0.050 |
| CP-*E. coli* to Both | 0.041 | 0.018 – 0.078 |
| CP-*Klebsiella* to Both | 0.045 | 0.019 – 0.087 |
| Both to CP-*E. coli* | 0.037 | 0.014 – 0.076 |
| Both to CP-*Klebsiella* | 0.040 | 0.018 – 0.073 |

**Supplementary Figure 2:**
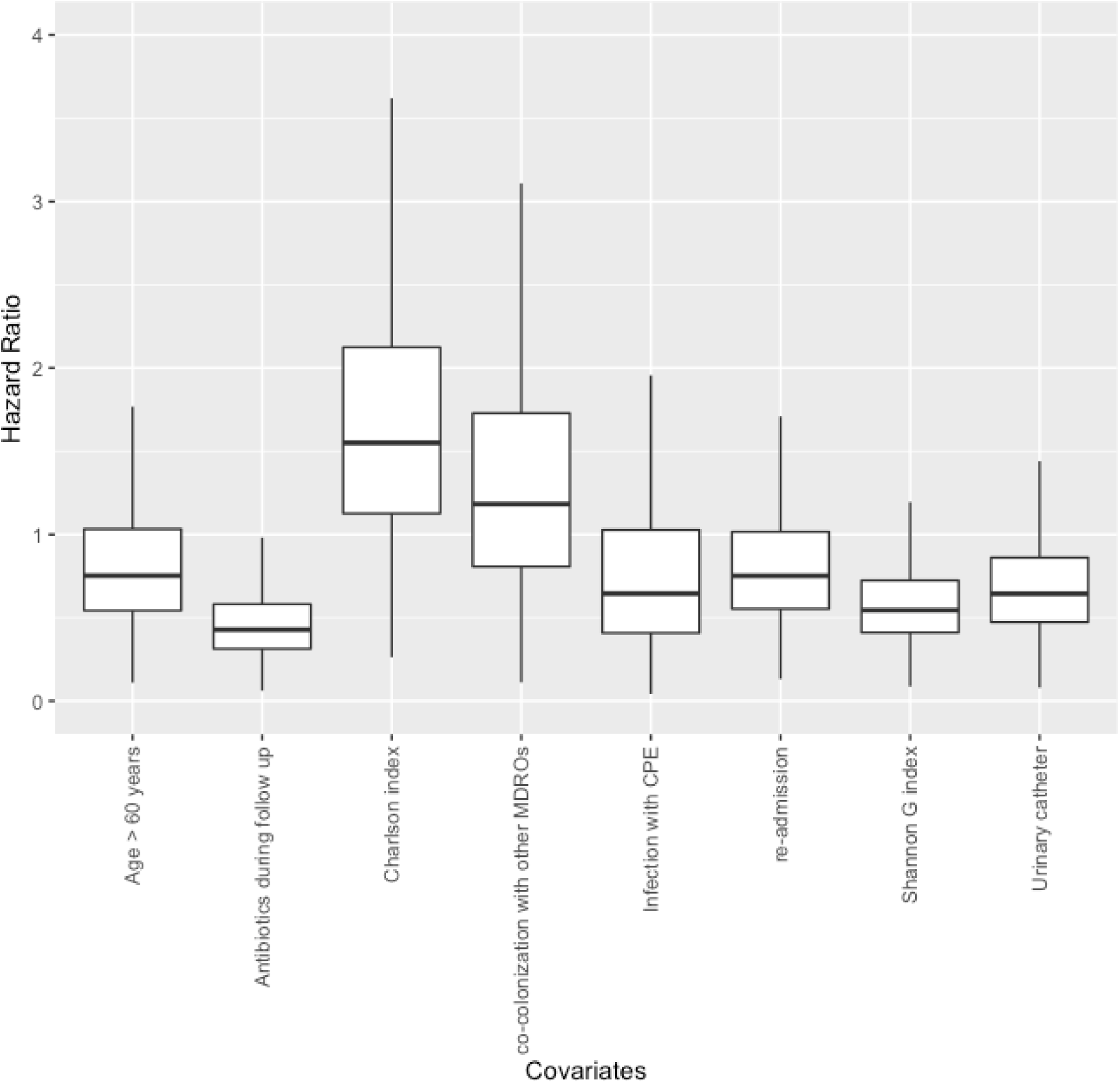
Associations of potential risk factors of CPE colonization and decolonization rate. MDRO: Multidrug resistant organisms including methicicllin-resistant *Staphylococcus aureus*, vancomycin-resistant *Enterococcus*, carbapenem-resistant *Enterobacteriaceae*, carbapenem-resistant *Acinetobacter baumanii* and carbapenem-resistant *Pseudomonas aeruginosa*.

**Supplementary Figure 3:**
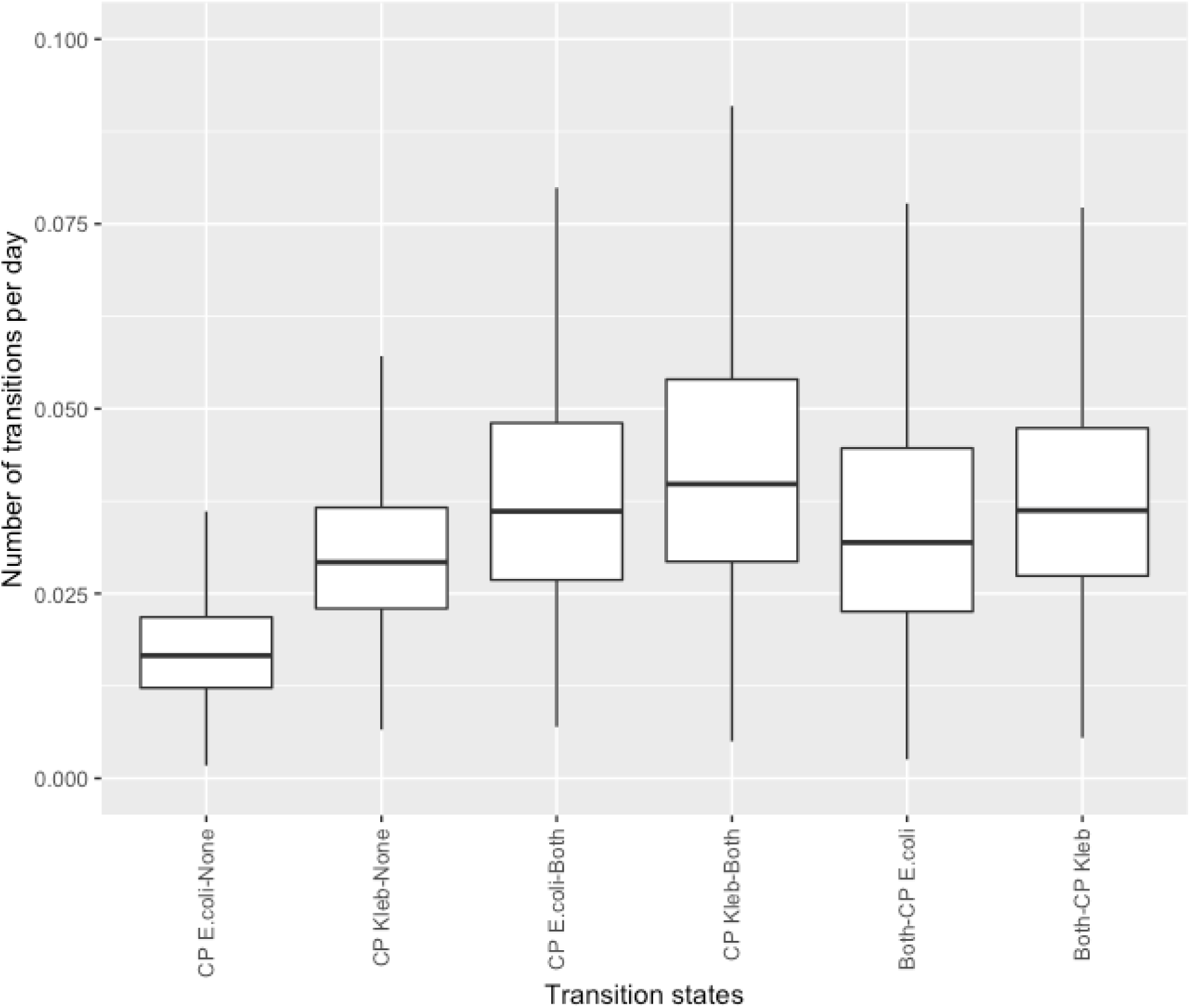
Rates of the four-state multistate model where CP-*E. coli and* CP-*Klebsiella pneumoniae* are separate states

## Notes

### Competing Interest Statement

The authors have declared no competing interest.

### Author Declarations

All relevant ethical guidelines have been followed and any necessary IRB and/or ethics committee approvals have been obtained.

Any clinical trials involved have been registered with an ICMJE-approved registry such as ClinicalTrials.gov and the trial ID is included in the manuscript.

## References

1. Logan LK, Weinstein RA. The Epidemiology of Carbapenem-Resistant Enterobacteriaceae: The Impact and Evolution of a Global Menace. J Infect Dis 2017; 215:S28–S36.

2. Magiorakos AP, Burns K, Rodríguez Baño J, et al. Infection prevention and control measures and tools for the prevention of entry of carbapenem-resistant into healthcare settings: guidance from the European Centre for Disease Prevention and Control. Antimicrob Resist Infect Control 2017; 6:113.

3. Centers for Disease Control and Prevention, US. Facility Guidance for Control of Carbapenem-resistant Enterobacteriaceae (CRE) – November 2015 Update CRE Toolkit. 2015. Available at: https://www.cdc.gov/hai/organisms/cre/cre-toolkit/index.html. Accessed 7 March 2019.

4. Teo JWP, La M-V, Krishnan P, Ang B, Jureen R, Lin RTP. Enterobacter cloacae producing an uncommon class A carbapenemase, IMI-1, from Singapore. J Med Microbiol 2013; 62:1086–1088.

5. Jari Oksanen, F. Guillaume Blanchet, Michael Friendly, Roeland Kindt, Pierre Legendre, Dan McGlinn, Peter R. Minchin, R. B. O’Hara, Gavin L. Simpson, Peter Solymos, M. Henry H. Stevens, Eduard Szoecs, Helene Wagner. Community Ecology Package. Available at: https://cran.r-project.org/web/packages/vegan/vegan.pdf. Accessed 7 March 2019.

6. R Core Team. R: A Language and Environment for Statistical Computing. 2018. Available at: https://www.R-project.org/.

7. Stan Development Team. RStan: the R interface to Stan. 2018; Available at: http://mc-stan.org/.

8. Tischendorf J, de Avila RA, Safdar N. Risk of infection following colonization with carbapenem-resistant Enterobactericeae: A systematic review. Am J Infect Control 2016; 44:539–543.

9. Bar-Yoseph H, Hussein K, Braun E, Paul M. Natural history and decolonization strategies for ESBL/carbapenem-resistant Enterobacteriaceae carriage: systematic review and meta-analysis. J Antimicrob Chemother 2016; 71:2729–2739.

10. Cerqueira GC, Earl AM, Ernst CM, et al. Multi-institute analysis of carbapenem resistance reveals remarkable diversity, unexplained mechanisms, and limited clonal outbreaks. Proc Natl Acad Sci U S A 2017; 114:1135–1140.

11. Mo Y, Seah I, Lye PSP, et al. Relating knowledge, attitude and practice of antibiotic use to extended-spectrum beta-lactamase-producing Enterobacteriaceae carriage: results of a cross-sectional community survey. BMJ Open 2019; 9:e023859.

12. Cassarly C, Martin RH, Chimowitz M, Peña EA, Ramakrishnan V, Palesch YY. Assessing Type I error and power of multistate Markov models for panel data—A simulation study. Communications in Statistics - Simulation and Computation. 2017; 46:7040–7061. Available at: http://dx.doi.org/10.1080/03610918.2016.1222425.

